# Baseline Characteristics and Outcomes of 180 Egyptian COVID-19 Patients Admitted to Quarantine Hospitals of Ain Shams University: A Retrospective Comparative Study

**DOI:** 10.1101/2021.06.11.21258743

**Authors:** Sara Ibrahim Taha, Sara Farid Samaan, Aalaa Kamal Shata, Shereen Atef Baioumy, Shaimaa Abdalaleem Abdalgeleel, Mariam Karam Youssef

## Abstract

**Background and study aim:** COVID-19 mortality, severity, and recovery are major global concerns, but they are still insufficiently understood, particularly in the Middle East. This study focused on evaluating if there was a link between COVID-19 patients’ clinical and laboratory findings at hospital admission and disease severity and mortality.

**Patients and methods:** A total of 180 adult Egyptian COVID-19 patients were included in this study then were categorized and compared.

**Results:** Of all, 27.8% had severe disease, and 13.9% died during their hospital stay. Diabetes (46.7%), hypertension (36.1%), and chronic obstructive pulmonary disease (COPD) (33.3%) were the most frequent associated co-morbidities. Severe patients and non-survivors were significantly older compared to their corresponding groups. Their neutrophil count, PCT, ESR, C-reactive protein (CRP), AST, ALT, LDH, D-dimer, and ferritin levels were significantly higher (P ≤ 0.05). In contrast, their absolute lymphocyte count was significantly lower (P ≤ 0.05). COPD (OR: 3.294; 95% CI: 1.199-9.053), diabetes (OR: 2.951; 95% CI:1.070- 8.137), ferritin ≥ 350 ng/mL (OR: 11.08; 95% CI: 2.796-41.551), AST ≥ 40 IU/L (OR: 3.07; 95% CI: 1.842-7.991), CT-scoring system (CT-SS) ≥ 17 (OR: 1.205; 95% CI: 1.089-1.334) and lymphocyte count < 1×103/µL (OR: 4.002; 95% CI: 1.537-10.421), were all linked to a higher risk of COVID-19 severity. While mortality was predicted by dyspnea (OR: 4.006; 95% CI: 1.045-15.359), CT-SS ≥ 17 (OR: 1.271; 95% CI: 1.091-1.482) and AST ≥ 40 IU/L (OR: 2.89; 95% CI: 1.091-7.661).

**Conclusions:** Clinical and laboratory data of COVID-19 patients at their hospital admission may aid in identifying early risk factors for severe illness and a high mortality rate.

## INTRODUCTION

Coronavirus disease 2019 (COVID-19) is an extremely infectious illness caused by the SARS-CoV-2 coronavirus, which was originally established in Wuhan, China, in early December 2019. Since then, the disease has rapidly spread to become a global epidemic [1]. The first COVID-19 case in Egypt was reported in February 2020, and since then, the number of cases has been dramatically rising. According to the Egyptian Ministry of Health and Population, more than 266,000 confirmed COVID-19 cases and almost 15,200 COVID-19 deaths throughout Egypt and its governorates as of June 5, 2021 [2].

COVID-19 infection is most commonly transmitted through respiratory droplets, as well as human-to-human contact [3]. It causes a wide range of symptoms that can lead to patient mortality, making it difficult to treat and control [4]. It also depletes hospital resources, such as ICU beds and mechanical ventilators, especially in low-resource countries [5]. So far, no specific antiviral therapy for COVID-19 is effective, and clinical management is based primarily on symptom control [6]. Several vaccinations began to emerge toward the end of 2020. According to the World Health Organization (WHO), there were 102 vaccines in clinical trials and 184 vaccines in preclinical trials as of May 28, 2021 [7].

This study aimed to understand more about COVID-19 baseline features and outcomes in Egyptian patients, which could aid in detecting early risk factors for poor prognosis and guiding proper patient management.

## PATIENTS AND METHODS

### Study subjects and settings

This retrospective study included 180 COVID-19 adult patients (age ≥ 18 years old) admitted from March 1 to April 30, 2021, to the Quarantine Hospitals of Ain-Shams University, Cairo, Egypt. Only confirmed SARS-CoV-2 patients were included in the study using RT–PCR tests on nasopharyngeal swabs, as per WHO guidelines [8]. Pregnant women, patients with missing data, and those who had hematological diseases or immunological disorders were excluded.

### Data collection

Baseline data including age, gender, presenting symptoms, co-morbidities and chronic diseases, chest computed tomography (CT), ICU admission necessity, length of hospital stay and outcome (survival or non-survival), as well as baseline routine laboratory test results (CBC with differential counts, CRP, ESR, ferritin, D-dimer, PCT, LDH, AST, and ALT) were acquired from medical records of all included patients.

### Ethical considerations

Before the study began, the Ain Shams University Faculty of Medicine Research Ethics Committee (REC) gave official consent. Because of the retrospective nature of this investigation, informed consent was not required. Data were acquired anonymously from hospital records and kept strictly confidential. They were only used for research purpose.

### Patient categorization

All included patients were categorized according to guidelines of the management protocol of COVID-19 patients released by Ain Shams University Hospitals [9]. Patients were considered to have severe/critical disease if one or more of the following were present: oxygen saturation ≤ 93% at rest; dyspnea with a respiratory rate ≥ 30 breath/min; arterial partial oxygen pressure (PaO2)/fraction of inspired oxygen (FiO2) ≤ 300 mmHg; respiratory failure with a need for mechanical ventilation; ICU admission need with shock, or organ failure syndrome. COVID-19 Patients with COVID-19 who did not match these criteria but had a positive COVID-19 nucleic acid test were classed as mild to moderately ill (non-severe).

### Calculation of CT scoring system

The level of involvement in each of the five lung lobes was visually examined, whether unilateral or bilateral; peripheral, central, or both; upper lobe predominance, lower lobe predominance, or both; and then scored on a scale of 0 to 5 (degree of involvement: 0%, less than 5%, 5–25%, 26–49%, 50–75%, and > 75%, respectively). The sum of the individual lobar grades was then used to create a score that ranged from 0 to 25 [10,11].

### Statistical analysis

The Statistical Package for Social Science (SPSS) was used to generate the results (version 26). Means and standard deviations (SD) or medians and ranges were used to describe quantitative numerical data. Frequencies and percentages were used to describe qualitative data. The Mann-Whitney –U test, the Chi-square test, or Fisher’s exact test were used to do comparisons. Stepwise logistic regression was used to estimate the risk for the relevant factors in the univariate analysis, and the odds ratio (OR) and its 95% confidence intervals (CI) were obtained. A p-value of ≤ 0.05 was judged significant.

## RESULTS

### Demographics and associated co-morbidities

A total of 180 COVID-19 patients were enrolled in this study. Their median (IQR) age was 48 years (15-94). Thirty-five percent of the patients were males, and 65% were females. The most common associated co-morbidities were diabetes mellitus (46.7%), hypertension (36.1%), and chronic obstructive pulmonary disease (COPD) (33.3%). Of all the studied population, 27.8% (n=50) had severe disease, and 13.9% (n=25) died during their hospital stay. Patients of these 2 subgroups were significantly older compared to non-severe patients (median: 60 years (IQR:22- 91) vs. 43 years (15-94); P=0.004) and survivors (57.0 years (27-80) vs 45.0 years (15-94); P=0.005), respectively. In terms of sex, differences between the studied subgroups were not significant. Demographics and associated co-morbidities are shown in **Table 1**.

**Table 1:** Sociodemographic characteristics and co-morbidities of all studied patients and according to COVID-19 severity and mortality.

|  |  | TOTAL | SEVERITY |  |  | MORTALITY |  |  |
| --- | --- | --- | --- | --- | --- | --- | --- | --- |
|  |  | N=180 | Non-severe<br>N=130 | Severe<br>N=50 | p-value | Survivors<br>N= 155 | Non-survivors<br>N= 25 | p-value |
| Age (years) | Median (IQR) | 48.0 (15-94) | 43 (15-94) | 60 (22-91) | **0.004** | 45.0 (15-94) | 57.0 (27-80) | **0.005** |
| Sex n., % | Male | 63 (35.0%) | 41 (31.5%) | 22 (44.0%) | 0.116 | 53 (34.2%) | 10 (40.0%) | 0.572 |
|  | Female | 117 (65.0%) | 89 (68.5%) | 28 (56.0%) |  | 102 (65.8%) | 15 (60.0%) |  |
| Co-morbidities<br>n., % | Diabetes | 84 (46.7%) | 49 (37.7%) | 35 (70.0%) | **≤0.001** | 67 (43.2%) | 17 (68.0%) | **0.021** |
|  | HTN | 65 (36.1%) | 36 (27.7%) | 29 (58.0%) | **≤0.001** | 49 (31.6%) | 16 (64.0%) | **0.002** |
|  | IHD | 25 (13.9%) | 14 (10.8%) | 11 (22.0%) | **0.051** | 18 (11.6%) | 7 (28.0%) | **0.028** |
|  | Stroke | 9 (5.0%) | 3 (2.3%) | 6 (12.0%) | **0.008** | 4 (2.6%) | 5 (20.0%) | **0.003** |
|  | CKD | 8(4.4%) | 1 (0.8%) | 7 (14.0%) | **≤0.001** | 6 (3.9%) | 2 (8.0%) | 0.353 |
|  | CLD | 36 (20.0%) | 20 (15.4%) | 16 (32.0%) | **0.013** | 31 (20.0%) | 5 (20.0%) | 1.000 |
|  | COPD | 60 (33.3%) | 35 (26.9%) | 25 (50.0%) | **0.003** | 52 (33.5%) | 8 (32.0%) | 0.879 |

*CKD: chronic kidney disease; CLD, chronic liver disease; COPD: chronic obstructive pulmonary disease; HTN, hypertension; IHD: ischemic heart disease. Significance (P-value) was set at $\leq 0.05$ .*

### Baseline clinical characteristics and patients’ outcomes

**Table 2** shows baseline clinical characteristics and outcomes of all studied patients and according to COVID-19 severity and mortality. The most frequent symptoms at admission were fatigue (90.6%), myalgia (75.0%), arthralgia (73.9%), fever (69.4%), dyspnea (69.4%) and cough (68.9%). Only dyspnea was significantly associated with COVID-19 in-hospital mortality (88.0% vs. 66.5%; P= 0.030). Patients with severe COVID-19 had significantly higher rates of thrombotic manifestations (38.0% vs. 22.3%; P=0.030), but no significant difference between non-survivors and survivors was found (36.0% vs. 25.2%; P=0.255). Regarding CT-SS, significantly higher scores were observed in severe patients (median: 21 (IQR: 5-23) vs. 15 (0- 24); P ≤ 0.001) and non-survivors (20 (0-24) vs. 7 (0-23); P ≤ 0.001) compared to their corresponding groups.

**Table 2:** Clinical characteristics and outcomes of all studied patients and according to COVID- 19 severity and mortality.

|  |  | TOTAL | SEVERITY |  |  | MORTALITY |  |  |
| --- | --- | --- | --- | --- | --- | --- | --- | --- |
|  |  |  | Non-severe | Severe | p-value | Survivors | Non-survivors | p-value |
|  |  | N=180 | N=130 | N=50 |  | N= 155 | N= 25 |  |
| Symptoms<br>n.,<br>(%) | Fatigue | 163<br>(90.6%) | 119 (91.5%) | 44 (88.0%) | 0.467 | 142<br>(91.6%) | 21<br>(84.0%) | 0.227 |
|  | Fever | 125<br>(69.4%) | 89 (68.5%) | 36 (72.0%) | 0.644 | 109<br>(70.3%) | 16<br>(64.0%) | 0.524 |
|  | Dyspnea | 125<br>(69.4%) | 88 (67.7%) | 37 (74.0%) | 0.411 | 103<br>(66.5%) | 22<br>(88.0%) | **0.030** |
|  | Arthralgia | 133<br>(73.9%) | 93<br>(71.5%) | 40<br>(80.0%) | 0.247 | 115<br>(74.2%) | 18<br>(72.0%) | 0.817 |
|  | Myalgia | 135<br>(75.0%) | 98<br>(75.4%) | 37<br>(74.0%) | 0.848 | 115<br>(74.2%) | 20<br>(80.0%) | 0.534 |
|  | Cough | 124<br>(68.9%) | 86<br>(66.2%) | 38<br>(76.0%) | 0.201 | 104<br>(67.1%) | 20<br>(80.0%) | 0.196 |
|  | Anosmia | 37<br>(20.6%) | 28<br>(21.5%) | 9<br>(18.0%) | 0.599 | 34<br>(21.9%) | 3<br>(12.0%) | 0.254 |
|  | Rhinitis | 18<br>(10.0%) | 16<br>(12.3%) | 2<br>(4.0%) | 0.096 | 17<br>(11.0%) | 1<br>(4.0%) | 0.281 |
|  | Headache | 65<br>(36.1%) | 46<br>(35.4%) | 19<br>(38.0%) | 0.744 | 55<br>(35.5%) | 10<br>(40.0%) | 0.663 |
|  | Chest tightness | 71<br>(39.4%) | 50<br>(38.5%) | 21<br>(42.0%) | 0.664 | 57<br>(36.8%) | 14<br>(56.0%) | 0.068 |
|  | Sore throat | 26<br>(14.4%) | 19<br>(14.6%) | 7<br>(14.0%) | 0.916 | 23<br>(14.8%) | 3<br>(12.0%) | 0.708 |
|  | Nausea | 22<br>(12.2%) | 18<br>(13.8%) | 4<br>(8.0%) | 0.283 | 20<br>(12.9%) | 2<br>(8.0%) | 0.487 |
|  | Diarrhea | 20<br>(11.1%) | 16<br>(12.3%) | 4<br>(8.0%) | 0.410 | 19<br>(12.3%) | 1<br>(4.0%) | 0.223 |
|  | Loss of taste | 12<br>(6.7%) | 11<br>(8.5%) | 1<br>(20.0%) | 0.120 | 11<br>(7.1%) | 1<br>(4.0%) | 0.565 |
|  | Thrombosis | 48<br>(26.7%) | 29<br>(22.3%) | 19<br>(38.0%) | **0.030** | 39<br>(25.2%) | 9<br>(36.0%) | 0.255 |
| CT-SS | Median (IQR) | 16.5 (0-24) | 15 (0-24) | 21 (5-23) | ≤**0.001** | 7 (0-23) | 20 (0-24) | ≤**0.001** |
| ICU admission | n., (%) | 55<br>(30.6%) | 7<br>(5.4%) | 48<br>(96.0%) | ≤**0.001** | 35<br>(22.6%) | 20<br>(80.0%) | ≤**0.001** |
| Hospital stay<br>(days) | Median (IQR) | 14 (3-29) | 10 (3-22) | 22 (10-29) | ≤**0.001** | 12 (3-29) | 22 (3-29) | **0.001** |
| In-hospital<br>mortality<br>n., (%) | Discharged<br>alive | 155<br>(86.1%) | 121<br>(93.1%) | 34<br>(68.0%) | ≤**0.001** |  |  |  |
|  | Died | 25<br>(13.9%) | 9<br>(6.9%) | 16<br>(32.0%) | ≤**0.001** |  |  |  |

CT-SS: CT-scoring system. Significance (P-value) was set at ≤0.05

In the 180 patients, the ICU admission rate was 30.6% with significant higher rates in severe patients and non-survivors compared to non-severe patients (96.0% vs. 5.4%; P ≤ 0.001) and survivors (80.0% vs. 22.6%; P ≤ 0.001). In-hospital mortality rates were significantly higher with severe compared to non-severe disease (32.0% vs. 6.9%; P ≤ 0.001). The median (IQR) duration of hospital stay of all studied patients was 14 days (3-29), with significantly longer durations in severe patients (22 days (10-29) vs. 10 days (3-22); P ≤ 0.001) and non-survivors (22 days (3-29) vs. 12 days (3-29); P=0.001) compared to their corresponding groups.

### Laboratory findings

Those with severe disease and non-survivors exhibited significantly higher (P ≤ 0.05) coagulation function (D-dimer), inflammation (PCT, ESR, CRP, and ferritin), liver dysfunction (AST and ALT), and tissue damage (LDH) compared to their corresponding groups. On the other hand, their CBCs showed significantly lower absolute lymphocyte counts (P ≤ 0.05) with significantly higher absolute neutrophil counts (P ≤ 0.05). There were no significant differences (P > 0.05) in hemoglobin level, total leukocyte, and platelet counts between the studied categories. **Table 3** shows the biochemical and hematological characteristics of the included patients.

**Table 3:** Baseline laboratory findings of all studied patients and according to COVID-19 severity and mortality

|  | TOTAL | SEVERITY |  |  | MORTALITY |  | p-value |
| --- | --- | --- | --- | --- | --- | --- | --- |
|  |  | Non-severe | Severe | p-value | Survivors | Non-survivors |  |
|  | n=180 | n=130 | n=50 |  | n= 155 | n= 25 |  |
| **TLC ($\times 10^3/\mu\text{l}$)** | 7.6 (0.1-79.0) | 7.5 (1.2-79.0) | 7.9 (0.1-24.0) | 0.856 | 7.6 (0.1-79.0) | 7.6 (1.2-21.6) | 0.849 |
| **NL ($\times 10^3/\mu\text{l}$)** | 2.5 (0.9-5.8) | 1.9 (0.9-4.2) | 2.9 (0.9-5.8) | **$\leq 0.001$** | 2.1 (0.9-4.2) | 2.6 (0.9-5.8) | **0.001** |
| **LYMPH ($\times 10^3/\mu\text{l}$)** | 1.2 (0.0-6.0) | 1.5 (0.1-6.6) | 0.9 (.0-3.6) | **$\leq 0.001$** | 1.3 (0.0-6.6) | 1.1 (0.1-2.4) | **0.005** |
| **HB (gm/dl)** | 11.4 (2.9-16.7) | 11.5(5.0-16.7) | 10.5 (2.9-15.0) | 0.280 | 11.2 (2.9-16.7) | 12.4 (6.6-15.2) | 0.104 |
| **PLT ($\times 10^3/\mu\text{l}$)** | 251.5 (6-716) | 254.0 (6-716) | 240.0 (11-650) | 0.627 | 256.0 (6-716) | 208.0 (6-485) | 0.124 |
| **Ferritin (ng/mL)** | 371.5 (16-4947) | 215.5 (16-2273) | 860.5 (130-4947) | **$\leq 0.001$** | 342.0(16-4947) | 878 (130-2577) | **$\leq 0.001$** |
| **PCT (ng/mL)** | 0.9 (0-5.9) | 0.8 (0.0-5.1) | 2.8 (0.4-5.9) | **$\leq 0.001$** | 0.9 (.0-5.9) | 2.5 (0.6-5.9) | **$\leq 0.001$** |
| **ESR (mm/h)** | 70.0 (12-210) | 57.0 (12-140) | 110.0 (49-210) | **$\leq 0.001$** | 65.0 (12-187) | 110 (30-210) | **$\leq 0.001$** |
| **CRP (mg/L)** | 48.0 (6.0-378.0) | 29.5 (6.0-372.0) | 102.5 (11-378.0) | **$\leq 0.001$** | 45.0 (6.0-378) | 91 (10-304) | **0.007** |
| **D-dimer (mg/L)** | 1.1 (0.1-12.5) | 0.8 (0.1-12.5) | 2.5 (0.7-12.2) | **$\leq 0.001$** | 0.9 (0.1-9.5) | 3.4 (0.6-12.5) | **$\leq 0.001$** |
| **LDH (IU/L)** | 222.0 (18-1889) | 189.0 (18-1889) | 392.0 (46-1121) | **$\leq 0.001$** | 207 (18-1889) | 388 (123 -1091) | **0.002** |
| **AST (IU/L)** | 26.0 (2-2044) | 25.0 (11-2044) | 45.0 (2-898) | **0.018** | 25 (2-2044) | 45 (12-898) | **0.002** |
| **ALT (IU/L)** | 30.0 (10-1344) | 25.0 (10-1344) | 45.0 (10-511) | **$\leq 0.001$** | 27 (10-1344) | 45 (18-511) | **0.001** |

*ALT: Alanine amino transferase; AST: Aspartate aminotransferase; CRP: C-reactive protein;*
*ESR: Erythrocyte sedimentation rate; HB: Hemoglobin; LDH: Lactate dehydrogenase; Lymph:*
*Lymphocytes; PCT: Procalcitonin; PLT: Platelets; NL: Neutrophils; TLC: Total leukocytic*
*count. Data presented as median (IQR). Significance (P-value) was set at $\leq 0.05$ .*

### Indicators of COVID-19 severity and in-hospital mortality

In **table 4**, multivariate logistic regression analysis was used to identify independent prognosis indicators associated with COVID-19 severity and in-hospital mortality. Predictors of COVID- 19 severity in the studied patients were presence of COPD (OR: 3.294; 95% CI: 1.199-9.053; P= 0.021) and diabetes mellitus (OR: 2.951; 95% CI:1.070-8.137; P= 0.037) as co-morbidities, abnormal high values of ferritin ≥ 350 ng/mL (OR: 11.08; 95% CI: 2.796-41.551; P= 0.001), AST ≥ 40 IU/L (OR: 3.07; 95% CI: 1.842-7.991; P= 0.021) and CT-SS ≥ 17 (OR: 1.205; 95% CI: 1.089-1.334; P ≤ 0.001), as well as low absolute lymphocyte count < 1×103/µL (OR: 4.002; 95% CI: 1.537-10.421; P= 0.005). Dyspnea as a presenting symptom (OR: 4.006; 95% CI: 1.045-15.359; P= 0.043), CT-SS ≥ 17 (OR: 1.271; 95% CI: 1.091-1.482; P= 0.002) and AST ≥ 40 IU/L (OR: 2.89; 95% CI: 1.091-7.661; P= 0.033) were also discovered to be independent predictors of COVID-19 in-hospital mortality.

**Table 4:** Multivariate logistic regression analysis of risk factors associated with COVID-19 severity and in-hospital mortality.

| Variables | SEVERITY |  |  |  |  |
| --- | --- | --- | --- | --- | --- |
|  | B | S.E. | P-value | Odds ratio | 95% CI |
| **Diabetes** | 1.082 | 0.518 | **0.037** | 2.951 | 1.070-8.137 |
| **COPD** | 1.192 | 0.516 | **0.021** | 3.294 | 1.199-9.053 |
| **CT-SS ($\geq 17$)** | 1.416 | 0.673 | $\leq 0.001$ | 1.205 | 1.089-1.334 |
| **Ferritin ($\geq 350$ ng/mL)** | 2.405 | 0.702 | **0.001** | 11.08 | 2.796-41.551 |
| **AST ($\geq 40$ IU/L)** | 1.124 | 0.487 | **0.021** | 3.07 | 1.842-7.991 |
| **Lymphocytes ($< 1 \times 10^3/\mu\text{L}$)** | 1.387 | 0.448 | **0.005** | 4.002 | 1.537-10.421 |
| MORTALITY |  |  |  |  |  |
| **Dyspnea** | 1.388 | 0.686 | **0.043** | 4.006 | 1.045-15.359 |
| **CT-SS ($\geq 17$)** | 2.023 | 0.792 | **0.002** | 1.271 | 1.091-1.482 |
| **AST ($\geq 40$ IU/L)** | 1.061 | 0.497 | **0.033** | 2.89 | 1.091-7.661 |

*AST: Aspartate aminotransferase; COPD: Chronic obstructive pulmonary disease; CT-SS: CT-scoring system. Significance (P-value) was set at $\leq 0.05$ .*

## DISCUSSION

In the current study, comprehensive data analyses of 180 hospitalized adult Egyptian COVID-19 patients were presented. The non-severe cases of COVID-19 (72.2%), as well as the survivors (86.1%), were by far the most prevalent in this study.

This study revealed that the severity and mortality of COVID-19 were significantly predominant in the older age groups. This could be due to the age-related reduction in cell-mediated and humoral immune functions, resulting from low-grade chronic inflammation [12], as well as the increased risk of multi-organ failure in elderly individuals due to their lower ability to correct for hypoxia [13]. Several studies have found that, when compared to young and middle-aged patients with COVID-19, older patients are more likely to proceed to severe disease with worse outcomes [14-16]. In contrast, Wang and colleagues couldn’t link COVID-19 severity or mortality to a certain age group [17].

COVID-19 has infected more males than females, according to numerous research [18,19]. The fact that women have fewer harmful behaviors than males, particularly smoking [20], suggests that gender may be a protective factor. Females’ innate and adaptive immune responses may be stronger than males’, making them more resistant to infections [21]. In terms of COVID-19 severity and morality, our research found no significant differences between male and female patients. Wang et al. and Fois et al., like us, discovered no gender differences in disease severity or mortality [12,22].

The most common chronic diseases among COVID-19 participants in our study were diabetes mellitus, hypertension, and COPD. These co-morbidities were discovered to be closely associated to the course of the disease. Furthermore, COPD and diabetes patients had probabilities of COVID-19 severity that were 3.294 and 2.951 times higher, respectively. These findings matched those of a meta-analysis study that identified diabetes mellitus and hypertension to be the most common underlying diseases among COVID-19 hospitalized patients [23]. Marhl et al. identified a higher risk of COVID-19 in diabetics because to the linked dysregulation of angiotensin-converting enzyme-2 (ACE-2), liver dysfunction, and chronic inflammation [24], which is consistent with our findings. In addition, the elevated risk of COVID-19 infection in patients with pre-existing cardiovascular illness may be due to a decrease in pro-inflammatory cytokines, which leads to weakened immune function [25]. In previous studies on MERS-CoV-2, it was discovered that the virus’s particular receptor, dipeptidyl peptidase-4, was expressed at a higher level in smokers and COPD patients [26]. Emami et al. suggested that COPD could be an underlying factor that makes patients more prone to COVID- 19 development [23], which is similar to our findings. Zhang et al., on the other hand, were unable to relate COVID-19 to COPD [27].

The most common symptoms found at the admission of our COVID-19 patients were; fatigue (90.6%), myalgia (75%), arthralgia (74%), fever (69.4%), dyspnea (69.4%), and cough (68.9%). In accordance with our results, Ghweli and coworkers found that malaise, fever, dry cough, and dyspnea were the most common manifestations of COVID-19 [14]. Dyspnea was found to be a significant prominent feature among the non-survivors in this study. Furthermore, the risk of disease mortality was 4.006 times higher in patients who presented with dyspnea than in those who did not. Similarly, Zheng et al. discovered a strong link between dyspnea and COVID-19 progression to death [28]. A meta-analysis research found similar results and suggested that dyspnea, rather than fever, should be used as a predictor of poor prognosis in COVID-19 patients [29].

Individuals with COVID-19 may have coagulation abnormalities, promoting a hypercoagulable state and resulting in an increased rate of thrombotic and thromboembolic events [30]. Using whole blood thromboelastography, Panigada et al. identified hypercoagulability features in COVID-19 patients, such as a decrease in time to fibrin formation, a decrease in time to clot formation, and an increase in clot strength [31]. In the context of these findings, a significant proportion of severe patients in our study were presented with thrombotic manifestations at admission to the hospital.

Severe patients tend to get priority hospitalization, with an increased need for oxygen supplementation, intensive care, and even mechanical ventilation [18]. Similarly, in our study, the length of hospital stay and the necessity for ICU admission were both considerably higher in severe patients and non-survivors.

During the early stages of COVID-19, when non-specific symptoms are present, total leukocyte and absolute lymphocyte counts are observed to be normal or slightly lower. Significant lymphopenia develops 1 to 2 weeks after the onset of the first symptoms, when the clinical signs of the disease worsen, accompanied by a significant increase in inflammatory mediators and cytokines in the bloodstream [32]. Several factors may play a role in COVID-19-related lymphopenia: the virus may directly infect lymphocytes, resulting in their lysis; markedly increased levels of interleukins may promote lymphocyte apoptosis; the cytokine storm may be linked to lymphoid organ atrophy; and metabolic acidosis, which is common in severe COVID- 19 patients, may suppress lymphocyte proliferation [32]. On the other hand, neutrophils are triggered by virus-related inflammatory factors produced by lymphocytes and endothelial cells, releasing reactive oxygen metabolites, neutrophil extracellular traps, and other cytotoxic mediators, which may suppress the virus [33]. Also, platelets may have an important role in the regulation of the virus-mediated inflammatory process [34]. The platelet count of most mild to moderate COVID19 patients may be normal or elevated, but it may be reduced in critically ill patients. In individuals with severe COVID19, thrombocytopenia may be caused by a decrease in platelet production, an increase in platelet breakdown, or a decrease in circulating platelets [35]. The current study revealed significantly lower median absolute lymphocyte counts and higher median absolute neutrophil counts, yet without neutrophilia, among severe patients and non-survivors. Also, lymphopenia < 1 × 103/µL was an independent predictor of disease severity (OR: 4.002). However, total leukocytic count, hemoglobin level, and platelet counts showed no significant differences as the disease progressed. In accordance with our results, lymphopenia has been recognized in many studies as an effective and reliable indicator of severity and mortality in COVID-19 patients [14,18,36]. Also, many studies reported that non-survivors presented with significantly higher neutrophil counts [4,36,37]. Moreover, Shang et al. found that hemoglobin levels were not influenced by the severity of the disease [38]. As opposed to our findings, Li et al. have found significant leucopenia and thrombocytopenia in severe patients [18]. In addition, Sulejmani et al. reported significant leukocytosis in severe COVID-19 patients, which could be related to the hyper-inflammatory state [39].

Viral infection has been linked to acute systemic inflammatory syndrome (ASIS), which includes fever and various organ dysfunction. Acute phase reactants are also produced by viral infection [40]. CRP is an inflammatory marker that aids in the resistance of invading pathogens in the host. Higher CRP levels have been connected to COVID19 complications such as heart injury, respiratory distress, and mortality. CRP measurement has been shown to be useful in determining the severity of COVID-19 patients [4]. Ferritin’s functions of iron binding and storage are also linked to the immunological and inflammatory responses to viral infection [41]. Procalcitonin (PCT) has recently been proposed as a useful inflammatory prognostic biomarker for identifying COVID-19 patients who are at high risk of clinical deterioration [42]. Inflammatory and tissue damage markers (CRP, ESR, ferritin, PCT, and LDH) were shown to be significantly greater in severe COVID-19 individuals and those who died from the disease. Ferritin levels of less than 350 ng/mL were also found to be an independent predictor of COVID- 19 severity, with an odds ratio of 11.08. These results matched those of a number of other studies [4,18,41,42].

Coagulation problems are rather common in COVID-19 patients with severe disease. The dynamics of D-dimer can indicate the severity and its elevated level is associated with adverse patients’ outcomes [14,30]. In accordance, D-dimer levels in our studied patients increased significantly as the disease worsened.

COVID-19 enters the human body through binding to the human ACE-2 receptor [43]. In 43.4 % of COVID-19 cases from Wuhan, Chen et al. reported a small increase in ALT and AST serum levels, which was the first report of hepatic dysfunction in SARS-CoV-2 infection [36]. Recently, a connection between abnormal liver biochemistry and COVID-19 severity was observed [18]. According to a study conducted in Shanghai [44], severe to critical COVID-19 cases had significantly higher serum ALT and AST levels than mild to moderate COVID-19 cases. In line with these findings, the odds ratios of AST ≥ 40 IU/L for COVID-19 severity and death were 3.076 and 2.890, respectively, in the current investigation, suggesting that it might be used as an independent predictor.

Patients with suspected SARS-CoV-2 infection can be isolated and treated in time for recovery, owing to the use of CT imaging in the diagnosis and grading of viral pneumonia [45]. Ground-glass opacities, consolidations, and Broncho-vascular thickening are characteristic chest CT findings in COVID-19 pneumonia. Atypical chest CT features also include masses, nodules, cavitation, lymphadenopathy, and pleural effusion [46]. The CT-scoring system (CT-SS), which shows how the severity of COVID-19 pneumonia affects dynamic changes in chest CT exams [46], was used to assess the extent of lung involvement in several studies [47, 48]. In this current study, the CT-SS showed significantly higher scores in the severe and non-survivor patients. Moreover, the odds of disease severity and mortality were 1.205 and 1.271 folds higher with CT- SS of ≥ 17, respectively. In a study by Hafez et al., the CT-SS 18/24 was regarded as the threshold value between mild and severe COVD-19 patients [46]. Furthermore, according to Francone et al., a CT-SS of 18 was significantly predictive of COVID-19 mortality [10].

There are certain limitations concerning this study that should be noted. First, it is a retrospective study that was performed in a limited hospital setting and included a relatively small sample size with disproportion in the different study groups. A large multi-center prospective observational study would be better to authenticate our findings. Another limitation is that all data included in this study were from the official records of the hospital; meanwhile, some patients have not been hospitalized for treatment because of the lack of awareness of disease severity, as well as the shortage of health care facilities. A larger study that includes out-patients isolated at home in addition to hospitalized in-patients would offer a more representative study population.

**In conclusion**, according to this study, COVID-19 infection was more aggressive in the elderly, diabetic, hypertensive, and COPD patients, as well as in those with low lymphocyte counts, high neutrophil counts, and high CRP, ESR, PCT, LDH, D-dimer, ferritin, AST, and ALT levels at the time of their admission to hospital. Hence, pretreatment clinical and laboratory data from COVID-19 patients at hospital admission may aid in identifying early risk factors for disease progression, as well as determining the most effective management plan.

## Data Availability

Data will be available upon request

## ACKNOWLEDGEMENT

The authors would thank all colleagues who contributed to conducting this study.

## ABBREVIATIONS

ACE-2: Angiotensin-converting enzyme-2
COPD: Chronic obstructive pulmonary disease
COVID-19: Coronavirus disease 2019
CT-SS: CT-scoring system
RT- PCR: Reverse transcription-polymerase chain reaction
SARS-CoV-2: Severe acute respiratory syndrome corona virus 2
WHO: World Health Organization.

